# Structure of Mental Disorders in Children in Outpatient Practice of a Specialized Mental Health Center in Tajikistan

**DOI:** 10.64898/2026.02.15.26346340

**Authors:** Mukadas F. Umarova, Mirsafo M. Mirsharofov

## Abstract

**Objective:** To analyze the structure of mental disorders in children in the outpatient practice of a specialized mental health center for optimization of care organization for this patient category.

**Methods:** A retrospective analysis of medical records of 23 children (out of 44 patients) at the Insight Mental Health Center (Dushanbe, Tajikistan) was conducted for the period from December 9, 2025, to January 8, 2026. Diagnosis was performed according to ICD-10 criteria using standardized instruments: M-CHAT-R, ADOS-2, and ADI-R for autism spectrum disorder (ASD); SNAP-IV for attention deficit hyperactivity disorder (ADHD); CGI; and pediatric versions of PHQ and GAD.

**Results:** Children accounted for 52% of all patients. Primary school-age children (7–12 years) predominated at 43.5%. Disorders of psychological development (F80–F89) dominated the nosological structure at 82.6%, with ASD comprising 56.5%. ADHD was diagnosed in 30.4% of cases. Comorbidity was registered in 47.7% of patients.

**Conclusion:** The structure of pediatric psychiatric pathology is characterized by a predominance of developmental disorders and high comorbidity levels, justifying the need for a multidisciplinary approach.

## Introduction

Mental disorders in children and adolescents represent a major global medical and social problem. According to the meta-analysis by Polanczyk et al., the worldwide prevalence of mental disorders among children and adolescents is 13.4%, corresponding to approximately 200 million young people globally [1]. According to the World Health Organization (WHO), approximately 50% of all mental disorders have their onset before the age of 14 years [2].

Autism spectrum disorder (ASD) occupies a prominent position in the structure of childhood psychopathology. According to the CDC (2023), ASD is identified in 1 in 36 children (2.8%) in the United States [3]. Lord et al. emphasize the importance of early ASD diagnosis using validated instruments such as ADOS-2 and ADI-R [4]. Early intervention significantly improves long-term outcomes [5].

Attention deficit hyperactivity disorder (ADHD) is one of the most prevalent neurodevelopmental disorders. According to the World Federation of ADHD (WFADHD) International Consensus Statement, the global prevalence of ADHD is 5–7% [6]. The high comorbidity rate of ADHD necessitates a comprehensive diagnostic approach [6,7].

Comorbidity of mental disorders in childhood presents a significant clinical challenge. Up to 40– 70% of children with ASD have co-occurring intellectual disability, and ASD-ADHD comorbidity reaches 30–50% [8,9]. Simonoff et al. demonstrated that 70% of children with ASD have at least one comorbid disorder [10].

In the Republic of Tajikistan, the child psychiatric care system is in a stage of active development [11]. Epidemiological data on the structure of childhood psychopathology in the country remain limited. The establishment of specialized mental health centers represents an important step in the development of child psychiatric services [12].

## Aim

To analyze the structure of mental disorders in children presenting to the Insight Mental Health Center (Dushanbe, Tajikistan) for optimization of care organization for this patient category.

## Materials and Methods

A retrospective analysis of medical records from the Insight Mental Health Center (Dushanbe) was conducted for the period from December 9, 2025, to January 8, 2026. During this period, 44 patients presented to the center, of whom 23 (52.3%) were children under 18 years of age.

Inclusion criteria were: age 0–17 years; established ICD-10 diagnosis; informed consent from a legal representative. Exclusion criteria were: incomplete documentation; refusal to participate.

Diagnosis was performed by a psychiatrist in accordance with ICD-10 criteria [13]. Standardized instruments were used to objectify the diagnostic process (Table 1).

**Table 1.**
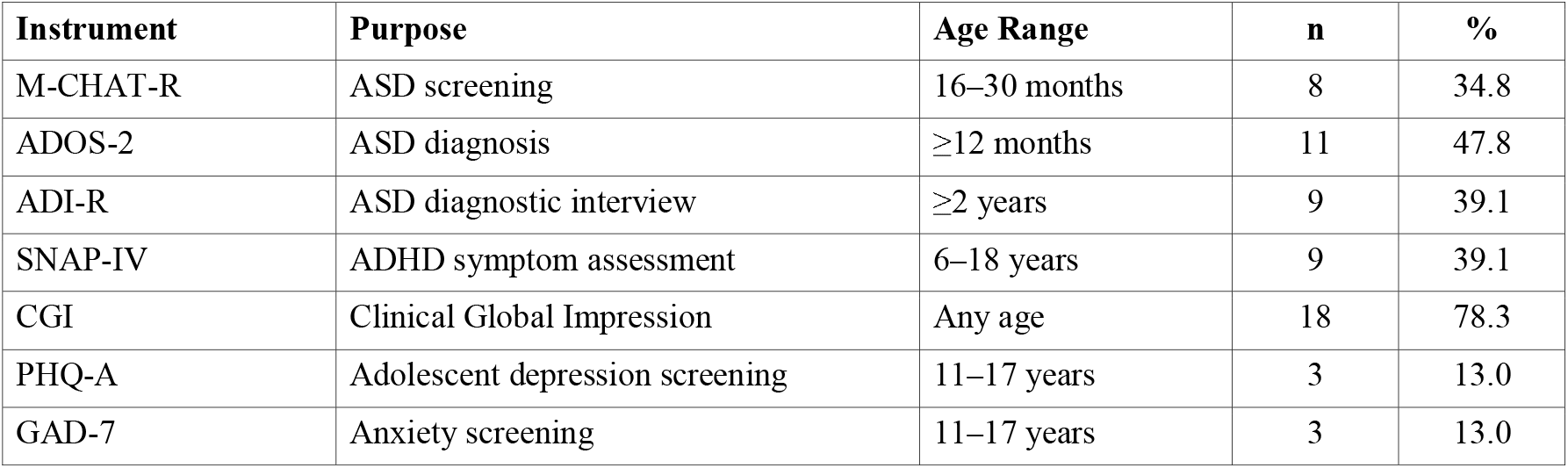
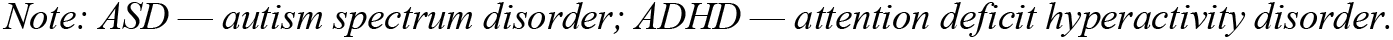
Standardized Diagnostic Instruments Used.

| Instrument | Purpose | Age Range | n | % |
| --- | --- | --- | --- | --- |
| M-CHAT-R | ASD screening | 16–30 months | 8 | 34.8 |
| ADOS-2 | ASD diagnosis | ≥12 months | 11 | 47.8 |
| ADI-R | ASD diagnostic interview | ≥2 years | 9 | 39.1 |
| SNAP-IV | ADHD symptom assessment | 6–18 years | 9 | 39.1 |
| CGI | Clinical Global Impression | Any age | 18 | 78.3 |
| PHQ-A | Adolescent depression screening | 11–17 years | 3 | 13.0 |
| GAD-7 | Anxiety screening | 11–17 years | 3 | 13.0 |

*Note: ASD — autism spectrum disorder; ADHD — attention deficit hyperactivity disorder.*

Statistical analysis included calculation of relative frequencies (%), means (M), and standard errors (±m). The study was conducted in accordance with the Declaration of Helsinki (2013).

## Results

### Sample characteristics

During the study period, children constituted 23 individuals (52.3% of all patients). The age and sex distribution is presented in Table 2.

**Table 2.** Age and Sex Distribution of the Pediatric Sample.

| Age Group | Males, n | Females, n | Total, n (%) |
| --- | --- | --- | --- |
| 0–3 years (early childhood) | 2 | 1 | 3 (13.0) |
| 4–6 years (preschool) | 5 | 2 | 7 (30.4) |
| 7–12 years (primary school) | 7 | 3 | 10 (43.5) |
| 13–17 years (adolescent) | 2 | 1 | 3 (13.0) |
| **Total** | **16 (69.6%)** | **7 (30.4%)** | **23 (100)** |

*Note: The predominant group consisted of preschool and primary school-age children (73.9%). Male-to-female ratio = 2.3:1.*

### Nosological structure

Analysis of the diagnostic structure revealed a predominance of disorders of psychological development (Table 3).

**Table 3.** Nosological Structure of Mental Disorders in Children.

| ICD-10 Code | Diagnosis | n | % |
| --- | --- | --- | --- |
| **F80–F89** | **Disorders of psychological development** | **19** | **82.6** |
| F84.0 | Childhood autism | 10 | 43.5 |
| F84.1 | Atypical autism | 3 | 13.0 |
| F80 | Specific developmental disorders of speech and language | 4 | 17.4 |
| F83 | Mixed specific developmental disorders | 2 | 8.7 |
| **F90–F98** | **Behavioural and emotional disorders with onset in childhood** | **11** | **47.8** |
| F90.0 | Attention deficit hyperactivity disorder | 7 | 30.4 |
| F91 | Conduct disorders | 2 | 8.7 |
| F93 | Emotional disorders with onset in childhood | 2 | 8.7 |
| **F70–F79** | **Intellectual disability** | **6** | **26.1** |
| F70 | Mild intellectual disability | 3 | 13.0 |
| F71 | Moderate intellectual disability | 2 | 8.7 |
| F72 | Severe intellectual disability | 1 | 4.3 |

*Note: Total exceeds 100% due to comorbid diagnoses in some patients.*

Comorbidity of mental disorders was registered in 11 patients (47.7%). The structure of comorbid combinations is presented in Table 4.

**Table 4.** Structure of Psychiatric Comorbidity.

| **Comorbid Combination** | **n** | **%** |
| --- | --- | --- |
| ASD + intellectual disability | 5 | 21.7 |
| ASD + ADHD | 3 | 13.0 |
| Speech disorders + developmental delay | 2 | 8.7 |
| ADHD + conduct disorder | 1 | 4.3 |
| **Total patients with comorbidity** | **11** | **47.7** |

### Diagnostic coverage

The mean number of assessments per patient was 2.8±0.3. Coverage with standardized diagnostics reached 77.3%. The most frequently used instruments were: CGI (78.3%), ADOS-2 (47.8%), and SNAP-IV (39.1%).

## Discussion

The study results demonstrate a high demand for child psychiatric services—children constituted more than half (52.3%) of all patients. This finding is consistent with global trends [1,2].

The predominance of preschool and primary school-age patients (73.9%) is explained by the most pronounced manifestation of neurodevelopmental disorder symptoms at this age, as well as increased cognitive demands at the onset of schooling [4,6].

A comparison of the obtained data with international studies is presented in Table 5.

**Table 5.** Comparison of Study Findings with International Data.

| Parameter | Current Study | International Data | Source |
| --- | --- | --- | --- |
| ASD proportion among referrals | 56.5% | 30–40% | Lord C. et al., 2018 [5] |
| ADHD proportion among referrals | 30.4% | 25–35% | Faraone S.V. et al., 2021 [6] |
| M:F ratio in NDD | 2.3:1 | 3–4:1 | Loomes R. et al., 2017 [14] |
| Comorbidity in ASD | 47.7% | 70% | Simonoff E. et al., 2008 [10] |
| ASD + intellectual disability | 21.7% | 31–40% | Lai M.C. et al., 2014 [8] |
| ASD + ADHD | 13.0% | 30–50% | Antshel K.M. et al., 2016 [9] |

*NDD — neurodevelopmental disorders.*

The higher proportion of ASD (56.5% vs. 30–40% in international studies) may reflect the center’s specialization as an institution with expertise in developmental disorder diagnosis. The lower comorbidity rate (47.7% vs. 70%) may be related to diagnostic practice characteristics and sample size [10].

The male-to-female ratio of 2.3:1 is consistent with international data on sex differences in neurodevelopmental disorders (3–4:1 for ASD, 2–3:1 for ADHD) [14].

The high coverage with standardized diagnostics (77.3%) and mean number of assessments (2.8±0.3) indicate adherence to evidence-based medicine principles in the center’s practice [15].

### Limitations

The study has several limitations, including a small sample size (n=23), a short observation period (1 month), and a single-center design. Future studies with larger samples will provide more representative data.

## Conclusions

1. Children constitute a significant proportion (52.3%) of patients at the specialized mental health center, reflecting the high demand for child psychiatric services.
2. The structure of pediatric psychiatric pathology is dominated by disorders of psychological development (82.6%), with ASD (56.5%) and ADHD (30.4%) being the leading diagnoses.
3. The high comorbidity rate (47.7%) justifies the need for a multidisciplinary approach involving allied specialists.
4. The use of standardized diagnostic instruments provides objectivity in diagnosis and creates a foundation for dynamic monitoring.

## Data Availability

All data produced in the present study are available upon reasonable request to the authors

## Declarations

### Ethics approval and consent to participate

The study was conducted in accordance with the Declaration of Helsinki of the World Medical Association (2013). Informed consent was obtained from the legal representatives of all study participants.

### Consent for publication

Not applicable.

### Availability of data and materials

The datasets used and analyzed during the current study are available from the corresponding author on reasonable request.

### Use of artificial intelligence

No artificial intelligence tools were used in the preparation of this article.

### Authors’ contributions

MFU: conceptualization, editing. MMM: data collection, writing, statistical analysis.

